# Metabolic syndrome and memory decline: evidence from a longitudinal aging cohort in rural South Africa

**DOI:** 10.1101/2025.10.28.25338982

**Authors:** Maria Mahuron Klein, Erika Beidelman, Thomas Gaziano, Chodziwadziwa Whiteson Kabudula, Molly Rosenberg

## Abstract

**Introduction:** Metabolic syndrome (MetS) is associated with increased risk of dementia in high-income countries. Given the different etiologic processes and population conditions driving MetS prevalence, it is unclear if MetS and dementia will show similar associations in low- and middle-income settings.

**Methods:** Mixed effects linear regression models were used to estimate the association between dichotomous MetS status and memory decline for individuals in the South African HAALSA Indepth cohort and by age and sex strata. An interaction term between MetS and time allowed the slope to vary by MetS status.

**Results:** MetS was associated with higher baseline memory scores (β = 0.07 SD units, 95% CI = 0.02, 0.13) and faster memory decline over time (β = −0.01 SD units/year, 95% CI = −0.02, 0.00).

**Discussion:** Our findings suggest that MetS status could be an important marker for identifying groups at higher risk of dementia in low-resource settings.

## 1. BACKGROUND

By 2050, more than 150 million people are projected to have aging-related dementias [1]. With no adequate dementia treatments, it is imperative to identify opportunities for dementia prevention through modifiable risk factors and determine those at greatest risk due to these factors [2]. One putative risk factor is metabolic syndrome (MetS) [3, 4], a condition defined by presence of at least three of the following: elevated waist circumference, elevated blood pressure, elevated blood glucose, elevated triglycerides, and reduced high density lipoprotein cholesterol (HDL-C) [5]. Globally, it is estimated that more than a billion people now meet the criteria for MetS [6].

The presence of MetS may indicate open pathways to increased dementia risk through insulin resistance, chronic inflammation, oxidative stress, and/or neurohormonal activation [7, 8]. Insulin resistance can cause inadequate glucose uptake to the brain [9], effectively starving it of energy needed for normal cognitive function. Chronic inflammation can cause atherosclerotic damage to blood vessels serving the brain [10]. Oxidative stress can disrupt the elimination of harmful free radical molecules produced as a result of the brain’s high metabolic activity [11]. Neurohormonal activation can release hormones such as leptin that trigger excess immune response while simultaneously suppressing other hormones such as adiponectin that protect against vessel damage [8]. The protective effects of sex-specific hormones such as estrogen on oxidative stress and inflammation imply the potential for differential associations between the component conditions comprising MetS and dementia for men and women, which has been noted by some systematic reviews [3, 12, 13].

More broadly, recent systematic reviews and meta-analyses generally confirm that MetS increases risk of dementia and predicts poorer performance on cognitive measures, although the findings are not wholly uniform [3, 7, 12–19]. Of the component conditions, elevated blood pressure and elevated blood glucose are most consistently associated with worse cognitive outcomes, but elevated waist circumference is found to have protective effects in some reviews [13, 14, 16]. Notably, studies from low- and middle-income countries (LMICs) are severely underrepresented in these reviews, and none included any study from sub-Saharan Africa.

Heterogenous effects of MetS component conditions on dementia risk suggest that populations with similar MetS prevalences may have differing MetS-associated dementia risk through different distributions in prevalence of component conditions. Component condition prevalences vary widely globally. Evidence from high-income countries highlight sedentary behavior, western dietary patterns, and smoking as the main drivers behind development of MetS component conditions [20]. However, high MetS prevalence estimates have also been observed in some LMICs still characterized by traditional diet and lifestyle [21, 22]. These prevalence estimates signal that the epidemiology of MetS in some LMICs is underpinned by non-western risk factors, resulting in a MetS composition distinct from that of high-income countries. Given the different etiologic processes and population conditions driving MetS prevalence in LMICs, it is unclear if MetS and dementia will show similar associations as those observed in high-income settings.

The existing evidence on the association between MetS and dementia in LMICs is largely cross-sectional [23–38]. Of the four longitudinal studies found, two found no association with MetS and two found an association with declining cognition [39–42]. No studies found a relationship with elevated waist circumference. The waist circumference component of MetS is of particular importance for sub-Saharan Africa, with debate on the appropriateness of the current European-based thresholds used to define elevated waist circumference in people of sub-Saharan descent [43–50]. Propensities for adiposity and insulin resistance differ between ancestral sub-Saharan and European populations [45, 46, 49].

The objective of this study is to determine the nature of the longitudinal relationship between MetS and episodic memory function in a rural, Black South African aging cohort [51]. From the limited evidence available on MetS and risk of dementia in other LMIC settings, we hypothesize that presence of MetS at baseline will be associated with faster memory decline over time.

## 2. METHODS

### 2.1 Study Population

The “Health and Ageing in Africa: Longitudinal Studies in South Africa” (HAALSA Indepth) is a cohort of men and women aged 40+ years, jointly led by the University of Witwatersrand and the Harvard TH Chan School of Public Health. Participants were randomly sampled in 2014 from the Agincourt Health and Socio-Demographic Surveillance System (HDSS), an annual census covering the entire (∼116,000) population of a ∼450 km^2^ rural area comprising 31 villages in Mpumalanga province in northeastern South Africa [52, 53].

The Agincourt HDSSS study area is located in a former ‘homeland’ region where Xitsonga-speaking Black South Africans were forcibly relocated from 1948-1993 during the apartheid era. During that period, the Black population in the region had poor access to education, poor public services, and high unemployment [54]. Historically, available jobs have been low-paying and in mining, farming, and domestic work [55]. While economic development in Agincourt has improved since the end of apartheid, unemployment remains high, and there remain gaps in basic services such as piped water and electricity [52]. Primary health care in the area is provided by six clinics, two health centers, and three district hospitals within 60 km [53].

Eligibility criteria for HAALSA Indepth included age 40 years and older as of 1 July 2014 and living permanently in the study area for 12 months before the 2013 Agincourt HDSS census. Of 12,875 eligible adults, 6281 were randomly selected using gender-specific sampling fractions to ensure a gender-balanced cohort [52]. The final baseline sample included 5059 adults. Retention in HAALSA Indepth is high, with 94% of surviving respondents in each of Wave 2 in 2018 and Wave 3 in 2021. Of the 5059 Wave 1 participants, we were able to ascertain MetS status for 4288 (85%).

### 2.2. Key Measures

#### 2.2.1. Exposure

The main exposure variable was MetS status at Wave 1 (2014/15). During Wave 1 interviews, trained field workers collected anthropometric measures and performed point-of-care blood tests via finger prick. We defined MetS using the 2009 harmonized criteria of the International Diabetes Federation and the American Heart Association/National Heart, Lung, and Blood Institute [5]. This definition requires the presence of at least three of the following five conditions: (i) elevated triglycerides or drug treatment for elevated triglycerides, (ii) reduced HDL-C or drug treatment for reduced HDL-C, (iii) elevated blood pressure or drug treatment for elevated blood pressure, (iv) elevated fasting glucose or drug treatment for elevated fasting glucose, or (v) elevated waist circumference [5]. Elevated triglycerides were defined as ≥1.7 mmol/L, reduced HDL-C as <1.0 mmol/L for men and < 1.3 mmol/L for women, elevated blood pressure as systolic pressure of ≥130 and/or diastolic pressure of ≥85 mm Hg, and elevated fasting glucose as ≥ 5.6 mmol/L. Just over 20% of our analytic sample had fasted for at least eight hours prior to glucose testing, so we set an additional threshold of 7.8 mmol/L for participants who were non-fasted or had missing fasting data to determine elevated blood glucose status [56]. Following the Harmonized definition’s recommendation to use ethnic-specific waist circumference thresholds, we defined ≥ 95.3 cm [49] and ≥ 92.0 cm [22, 45] as elevated for men and women respectively. To avoid possible overestimation of elevated waist circumference, we selected the highest thresholds reported from the five largest African optimization studies we located [22, 45, 46, 48, 49].

Each component condition was assigned a value of 1 if it met the Harmonized definition threshold and a value of 0 if it did not. Presence of MetS was defined as having a value of 1 for three or more component conditions, whereas no MetS was defined as having a value of 0 for three or more component conditions. This MetS dichotomization method has been employed by other South African MetS studies and allowed us to assign MetS status even when up to two component condition measurements were missing [22, 47, 48, 57].

#### 2.2.2. Outcome

The outcome was episodic memory score decline between Waves 1 and 3. A brief cognitive battery harmonized with the battery used in the US Health and Retirement Study was administered at each in-person HAALSA Indepth interview [52]. Memory scores were calculated based on immediate and delayed word recall trials of a 10-word list read out loud by the interviewer at each HAALSA Indepth wave. To account for differences in the administration of the word recall trials across waves, these memory scores were constructed using confirmatory factor analysis, then z-standardized [58]. This method has been successfully implemented in other HAALSA Indepth studies utilizing memory scores as an outcome [59, 60].

#### 2.2.3. Covariates

Other key measures were included to characterize the study population, control for potential confounding, and assess potential effect modification. These covariates were: age, considered continuously and categorically (ages 40-62, heretofore called mid-life and 62+, heretofore called later-life), sex (male vs. female), having children (any children vs. no children), literacy (able to read and/or write vs. not able to read and write), educational attainment (none/some primary vs. some secondary and higher), household consumption quintile (one lowest, five highest), marital status (partnered vs. not partnered), self-reported HIV status (HIV-positive vs. HIV-negative), and country of birth (South Africa vs. other). Household consumption quintile was determined by totaling all reported annualized household expenditures and food consumption values, dividing by the number of persons reported in the household, and assigning a value of one to five based on the percentile of the per capita value.

### 2.3. Statistical Analysis

We fit mixed effects linear regression models to estimate the association between the dichotomous MetS status and memory scores over time, as well as models for each component condition and memory scores over time. Models were specified for both the total sample and for men and women separately. We stratified the models by age category (mid-life and later-life) to assess possible differential associations for younger and older participants.

All mixed effects models were fit with a random slope for time, random intercept for the individual, and inverse probability weights for attrition and mortality over time. Inclusion of the random effects allowed us to account for correlation between an individual’s observations over time and individuals within the same household. We accounted for practice effects (i.e., improvements in test scores resulting from repeat testing) using dummy variables indicating first recorded memory score [61]. We modeled memory decline with an interaction between MetS (or component condition) and time to allow the slope to vary by MetS/component condition status. Additional covariates specified in the adjusted models included age, having children, literacy, education level, household consumption, partnered status, self-reported HIV status, and country of birth.

We performed two sensitivity analyses to test the robustness of our results to our analytic choices of waist circumference thresholds and treatment of fasting status in determination of elevated blood glucose. For the first sensitivity analysis, we redetermined MetS status using the standard Europid thresholds for waist circumference (94.0 cm for men and 80.0 cm for women) [5]. For the second sensitivity analysis, we excluded 52 participants with missing fasting data from determination of elevated blood glucose status instead of assuming they were non-fasted.

Significance levels for all test statistics were set at 0.05. Statistical analyses were performed in RStudio version 2023.12.1+402 [62]. Log-binomial generalized linear models were estimated using the ‘stats’ base R package. Mixed effects models were estimated with the ‘lme4’ package [63].

## 3. RESULTS

### 3.1. Sample Characteristics

Among the 5059 study participants, the median age was 62 years (Range: 40-112 years) (Table 1). Among MetS status groups there were significant differences in median age, sex, country of birth, having children, and reported HIV status. Those with MetS were more likely to be female, older, born in South Africa, and have children. Those without MetS were more likely to report living with HIV.

**Table 1.**
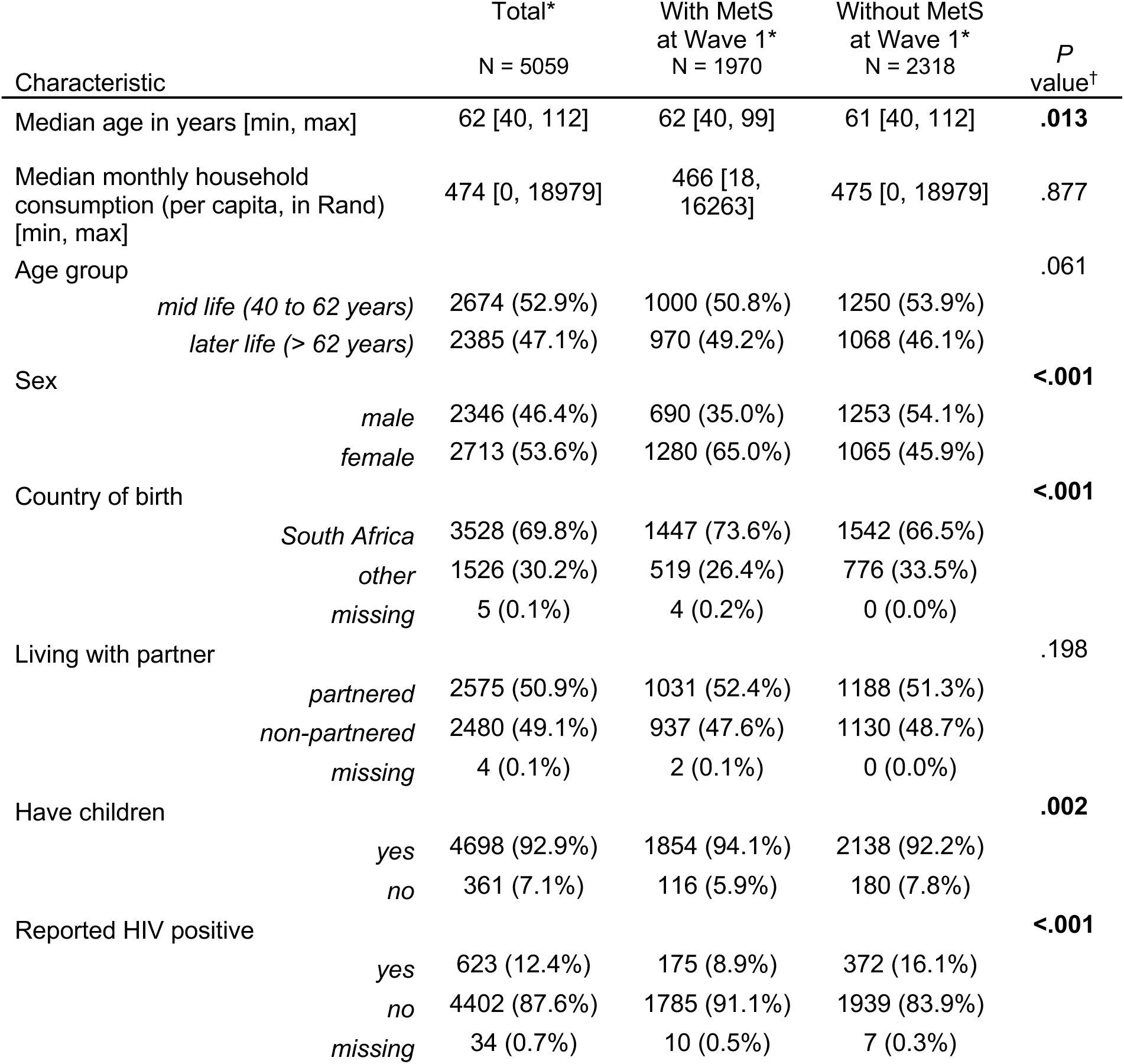
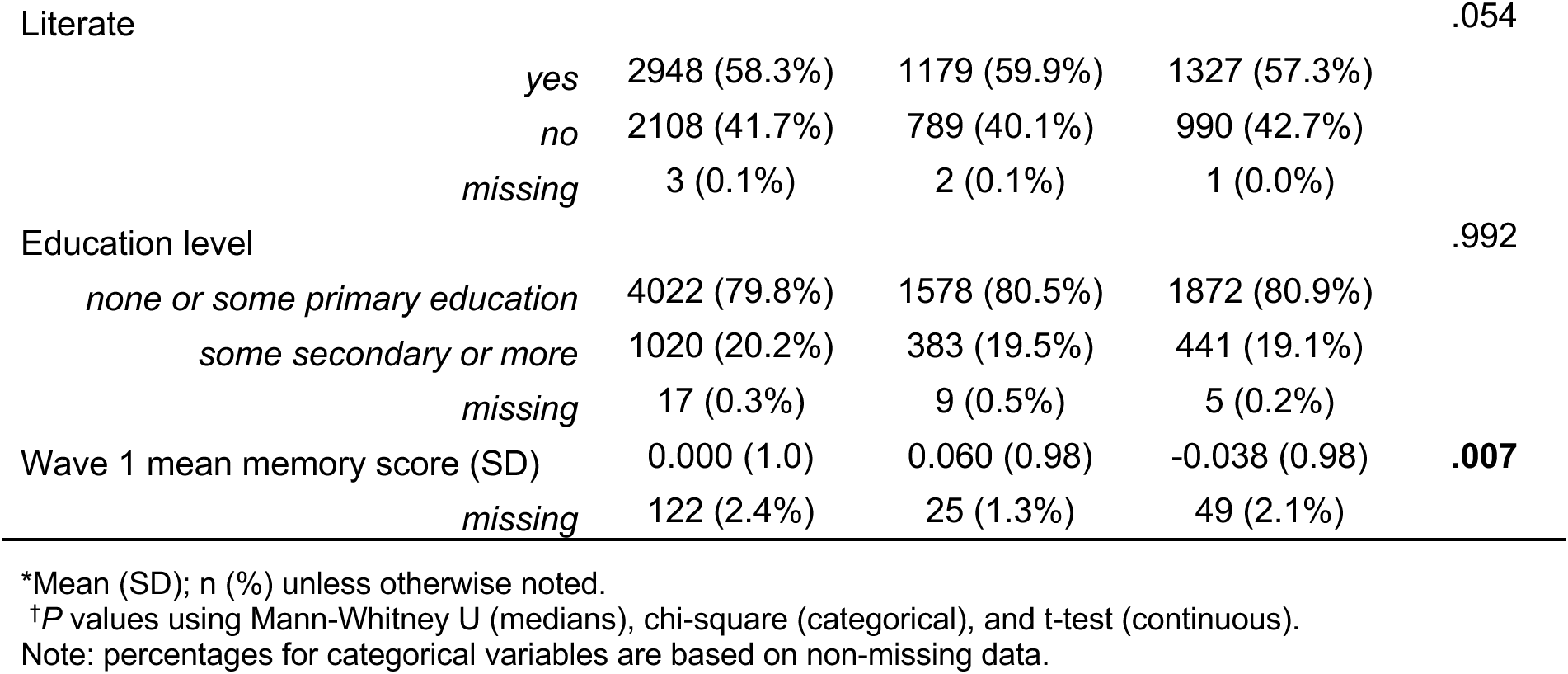
Sociodemographic characteristics by metabolic syndrome (MetS) status: HAALSA Indepth, 2014/15.

### 3.2. MetS Status and Memory Score

Having MetS was associated with higher memory scores at baseline and with faster memory decline over time (Figure 1). In the full population, those with MetS had higher baseline memory scores by 0.07 SD units (95% CI = 0.02, 0.13) and faster memory decline over time (β = −0.01 SD units per year, 95% CI = −0.02, 0.00). The direction and magnitude of these estimates did not differ by sex or age categories (Table 2).

**Figure 1.**
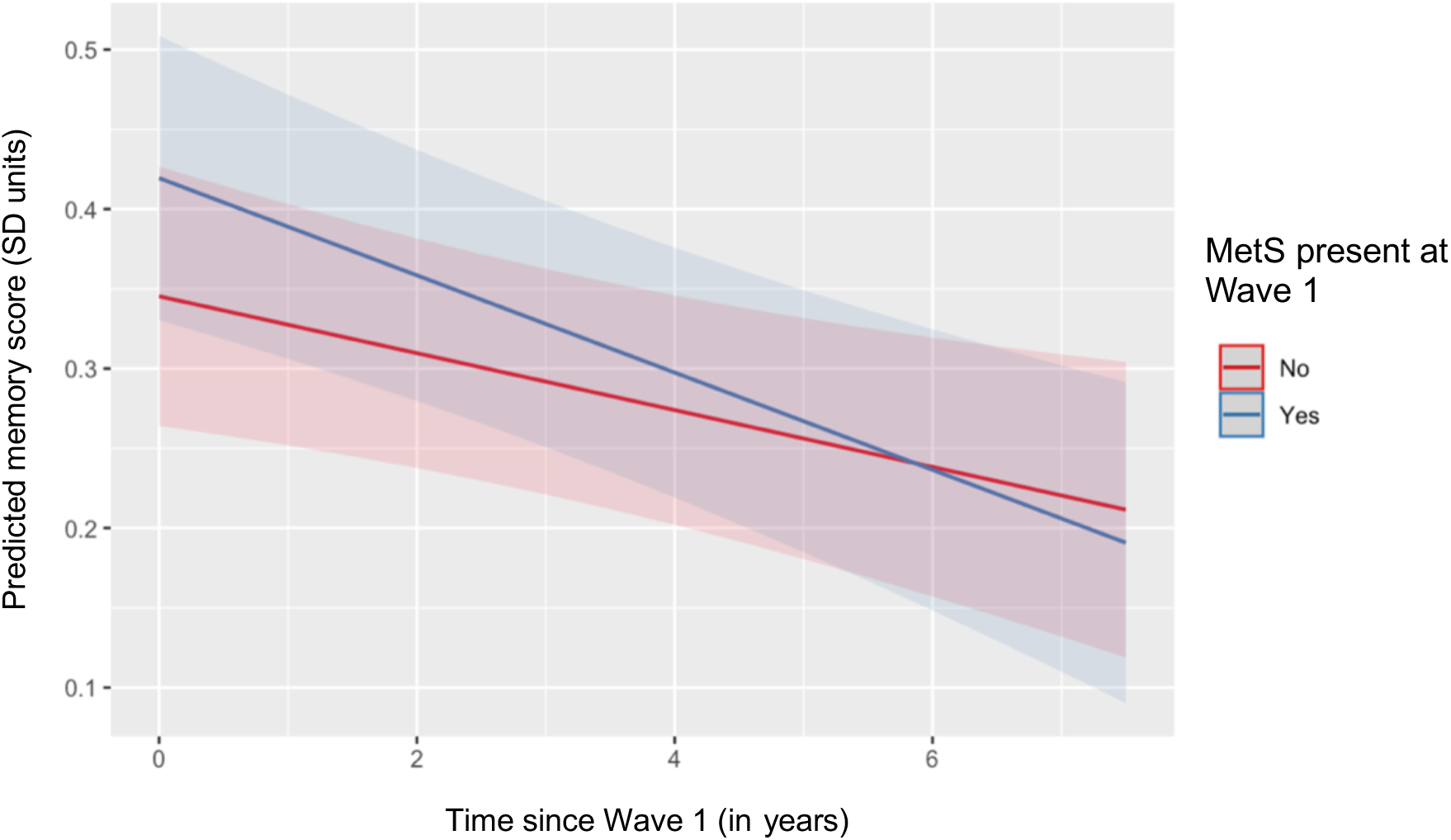
Predicted memory scores by metabolic syndrome (MetS) status for HAALSA Indepth participants, 2014/15-2021/22.

**Table 2.** Effect of metabolic syndrome (MetS) on z-standardized memory scores for HAALSA Indepth participants: 2014/15-2021/22.

|  |  | Baseline memory score |  |  | Memory score decline |  |  |
| --- | --- | --- | --- | --- | --- | --- | --- |
| | | $\beta$ | 95% CI | P value | $\beta$ | 95% CI | P value |
| Full population |  |  |  |  |  |  |  |
|  | Unadjusted | **0.08** | **(0.02, 0.14)** | **0.008** | **-0.01** | **(-0.03, 0.00)** | **0.015** |
|  | Adjusted* | **0.07** | **(0.02, 0.13)** | **0.009** | **-0.01** | **(-0.02, 0.00)** | **0.030** |
| By Sex |  |  |  |  |  |  |  |
|  | Men | 0.07 | (-0.02, 0.16) | 0.128 | -0.01 | (-0.03, 0.01) | 0.348 |
|  | Women | 0.07 | (0.00, 0.14) | 0.061 | -0.01 | (-0.03, 0.00) | 0.086 |
| By Age* |  |  |  |  |  |  |  |
|  | Mid-life | 0.07 | (-0.01, 0.15) | 0.076 | -0.01 | (-0.03, 0.00) | 0.092 |
|  | Later-life | 0.06 | (-0.02, 0.14) | 0.115 | -0.01 | (-0.03, 0.01) | 0.173 |
| By Sex and Age† |  |  |  |  |  |  |  |
|  | Mid-life men | 0.08 | (-0.06, 0.22) | 0.270 | -0.02 | (-0.05, 0.01) | 0.218 |
|  | Later-life men | 0.07 | (-0.06, 0.19) | 0.289 | 0.00 | (-0.03, 0.02) | 0.846 |
|  | Mid-life women | 0.07 | (-0.03, 0.16) | 0.172 | -0.01 | (-0.03, 0.01) | 0.296 |
|  | Later-life women | 0.05 | (-0.05, 0.15) | 0.366 | -0.01 | (-0.04, 0.01) | 0.170 |

\*Adjusted for sex, continuous age, having children, literacy, education level, consumption quintile, marital status, self-reported HIV status, and country of birth.
†Adjusted for continuous age, having children, literacy, education level, consumption quintile, marital status, self-reported HIV status, and country of birth.
Note: Effect estimate in standard deviation units.

### 3.3. Component Conditions and Memory Score

Elevated waist circumference had the strongest association with memory scores among all the MetS component conditions (Table 3). Baseline memory scores were higher and memory score decline was faster for participants with elevated waist circumference compared to those without (Figure 2). Higher baseline scores and faster rates of decline were also observed for elevated triglycerides and elevated blood pressure. The rate of decline indicates that, even given the higher baseline, memory scores were lower for those with elevated triglycerides and elevated blood pressure by the end of the follow up period. No statistically significant relationships were observed between memory score and reduced HDL-C or elevated blood glucose. Across age and sex strata, baseline memory scores and differences in rate of memory decline were generally similar in magnitude within each component condition. The one exception was for elevated blood pressure, for which the difference in rate of memory decline between those with and without elevated blood pressure was more extreme among later-life participants than mid-life (Supplementary Table 1). Due to rounding, the point estimate confidence intervals for difference in rate of decline in these age groupings appear to overlap but significance testing indicates a statistical difference between them (−0.03, 95% CI: −0.05, −0.00, *P* value = .034).

**Figure 2.**
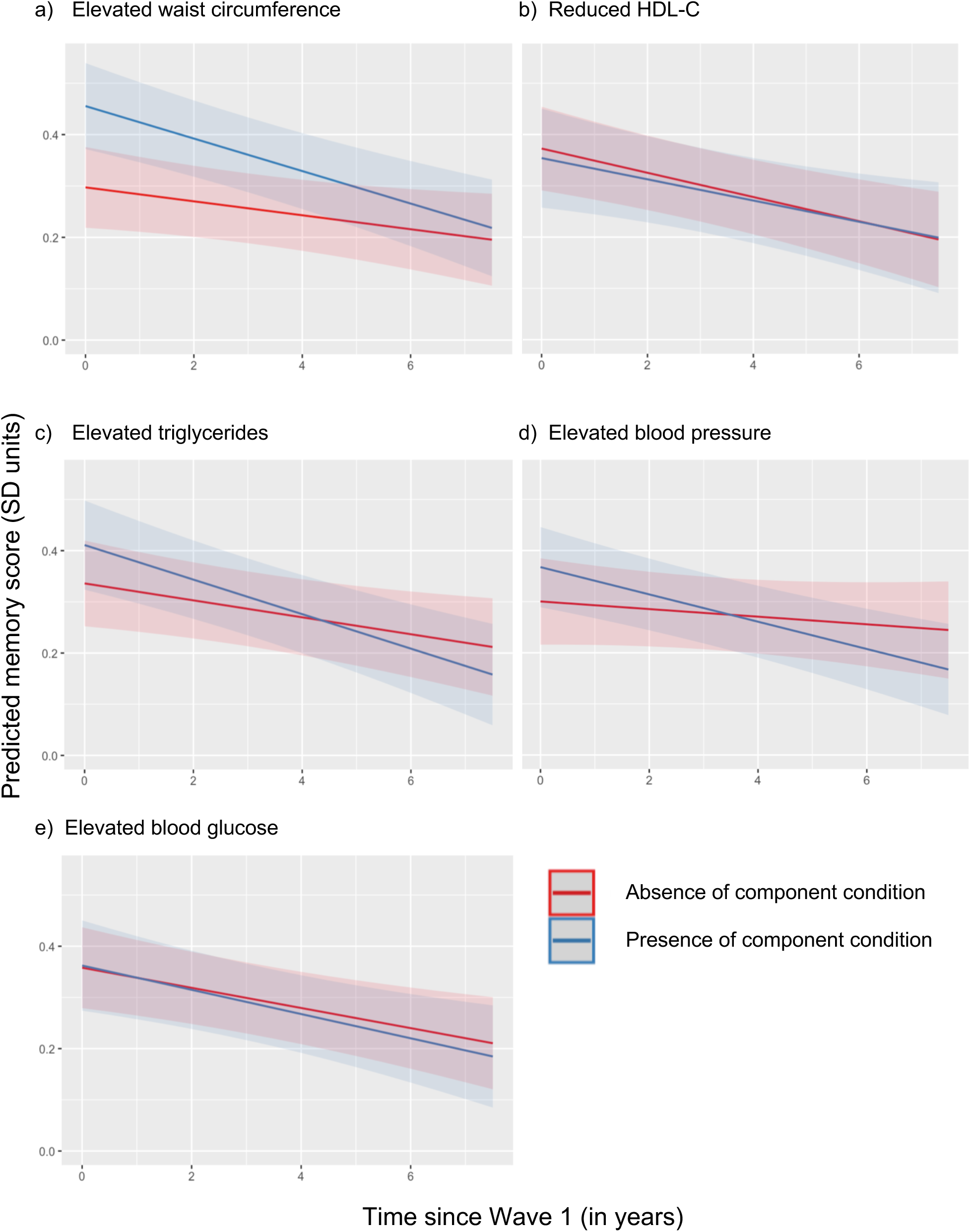
Predicted memory scores by metabolic syndrome (MetS) component conditions for HAALSA Indepth participants, 2014/15-2021/22.

**Table 3.**
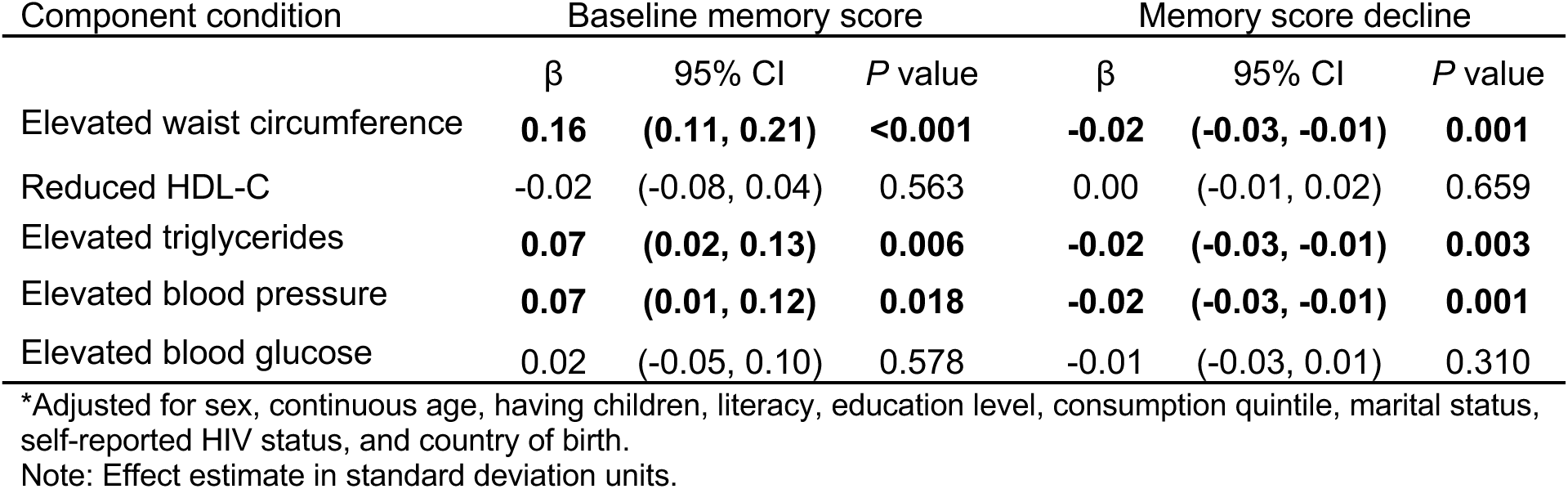
Adjusted* effect of metabolic syndrome (MetS) component conditions on z-standardized memory scores for HAALSA Indepth participants: 2014/15-2021/22.

### 3.4. Sensitivity Analyses

When using the Europid-defined MetS status, the associations between MetS status/elevated waist circumference and baseline memory score and memory decline over time maintain the same direction and generally similar magnitudes among all strata (Supplementary Table 2). In the main analysis, mid-life associations were marginally significant and reach statistical significance in the sensitivity analysis, with similar point estimates. When excluding those with missing fasting data from determination of elevated blood glucose, the association between MetS and memory score likewise maintain the same directions and similar point estimate and confidence interval magnitudes (Supplementary Table 3).

## 4. DISCUSSION

In this study, we characterized the longitudinal relationship between MetS and memory decline in an aging cohort of rural, Black South Africans in a low-income setting. We observed a MetS prevalence of 36% among our sample of adults aged 40+ years. Presence of MetS was associated with higher baseline memory scores and faster memory decline over time. Among the individual component conditions, elevated waist circumference had the strongest association with memory scores. In terms of memory decline and the predicted memory score by the end of follow-up, presence of MetS appears to predict memory decline similarly to two of its component conditions, elevated triglycerides and elevated blood pressure.

To our knowledge, this is the first study investigating the relationship between MetS and longitudinal cognitive outcomes in sub-Saharan Africa. The most recent African meta-analyses cast doubt that criteria for MetS developed using Western populations provide the same predictive value for adverse health outcomes such as dementia in African populations [43, 44]. However, consistent with our hypothesis and with evidence from high-income settings, we found evidence that MetS is associated with faster memory decline. Our findings also align with work from high-income countries in identifying an association between elevated blood pressure and poorer cognitive outcomes. This was especially true for later-life men with elevated blood pressure, who showed the fastest rate of decline associated with the presence of any individual component condition.

As reported in some literature from high-income settings, we found a protective effect of elevated waist circumference on memory score at baseline [13, 14, 16]. However, the faster rate of decline suggests this may be a waning effect. This result could have been a function of our analytic choice of sub-Saharan-specific waist circumference thresholds, although a sensitivity analysis using Europid thresholds showed robustness of the results to this specification. It is also possible that the faster decline in conjunction with the higher baseline score is a function of statistical phenomena such as regression to the mean. It is worth noting that although the rate of memory score decline was faster, the baseline memory score was high enough that the decline did not fully offset the difference; people with elevated waist circumference still had higher memory scores by the end of the approximately 7-year follow up period.

Our study did not find the association between elevated blood glucose and cognitive decline generally found in high-income settings. This may be a function of our treatment of fasting status, in which we required any participant who was non-fasted or had missing fasting data to reach a higher glucose threshold to be classified as having elevated blood glucose. However, a sensitivity analysis excluding blood glucose status from determination of MetS for any participants with missing fasting data yielded similar results to the main analysis. Additionally, single blood glucose readings are sensitive to external factors such as quality of prior night’s sleep, time of day, and recency of physical activity [64]. More robust measures of impaired glucose metabolism such as HbA1c may yield different associations.

The higher baseline memory score associated with presence of MetS is a departure from evidence in high-income regions [13], but evidence available from LMICs is mixed. MetS has been associated with increased odds of cognitive impairment, lower CERAD Neuropsychological Battery and Mini-Mental State Examination (MMSE) scores, poorer word recall performance, and decreased digit symbol substitution test scores in some LMIC studies [27, 28, 30, 31, 34]. Others, including our own study, report no or even positive associations between presence of MetS and cross-sectional cognitive outcomes [23, 24, 29, 38]. No association was found between MetS and presence of a motoric cognitive risk, a pre-dementia state characterized by reduced walking speed and subjective cognitive concerns [39]. Positive associations have been reported between MetS and MMSE scores among the oldest old [34] as well as higher point estimates for episodic memory, executive function, orientation, registration, attention, calculation, verbal fluency, and constructional praxis domain scores, although the estimates for individual cognitive domains were not generally significant and survival bias was noted as a possible explanation [28, 30, 40].

Our findings of higher memory score at baseline for those with MetS could be explained by higher prevalence of nutritional insecurity in LMIC regions [65]. Food insecurity is not only the absence of food; it is also defined as an unstable household food supply or overconsumption of cheap, energy-dense foods instead of more expensive, nutrient-dense alternatives [66]. In South Africa, for example, diets are generally low in fruit and vegetable consumption [67] and high in starchy carbohydrates associated with obesity, high triglycerides, and reduced HDL-C [68]. Socioeconomic factors that are associated with better cognitive outcomes such as education, income, and consumption are also associated with increased food purchasing power [69]. In low-resource settings, these socioeconomic factors may allow for expanded food purchasing and consumption, but not nutritional security in terms of access to food more conducive to cardiometabolic health. Our observation of the strongest memory score associations for those with elevated waist circumference lends to support to this explanation.

Though we adjusted for education level and per capita household consumption, the observed positive relationship between MetS and baseline memory score may be due in part to residual confounding from unmeasured factors related to socioeconomic position.

Strengths of this analysis include an exposure constructed using point-of-care biomarker measurements rather than self-reported status, reducing misclassification bias. Use of sub-Saharan-specific waist circumference thresholds rather than European-derived thresholds also increases the likelihood of correctly classifying participants at increased risk. The z-standardized memory measure is also a strength, as it was constructed from recall tests which measure episodic memory [70]. Episodic memory is typically the first of memory systems to decline with dementia, enabling the earliest detection of longitudinal memory changes [71, 72]. Additionally, use of a population-based study cohort with a high retention rate and rich sociodemographic data increases the likelihood that our findings are representative of the source population of older adults in rural South Africa.

Some aspects of our study warrant careful interpretation of our findings. Informative censoring due to study drop-out or mortality is possible. Even given the relatively high cohort retention rate, participants with the most severe manifestations of MetS or memory decline may not have recorded observations at all follow up times. To address this possibility, we used mortality and attrition weights over the follow-up to minimize the likelihood of biased results due to differential death or drop out [61]. However, pre-baseline mortality that occurred prior to cohort enrollment will still be present and may introduce selection bias into our study population. Additionally, our exposure is sensitive to the accuracy of the point-of-care measurements. Inaccuracy in measurement, as well as inaccuracy introduced by our analytic choice to dichotomize continuous outcomes, could lead to misclassification bias. We performed a sensitivity analysis using a different dichotomization threshold for elevated waist circumference to test the robustness of our results to changes in MetS definition and found very similar point estimates and confidence intervals, alleviating some concern about misclassification. Finally, our analyses by age and sex may not be sufficiently powered to detect significant differences.

## 5. CONCLUSION

Our findings indicate that MetS is associated with faster memory decline in rural, aging Black South Africans. As such, using MetS could prove useful to predict dementia risk in this population. Unlike some other dementia risk identifiers, MetS status can be established without genetic testing, advanced laboratory work, or expensive imaging. Thus, MetS status could be an important marker for identifying groups at higher risk of dementia in low-resource settings. Future studies should explore associations using different operationalizations of MetS, specific component condition combinations, additional cognitive domains, incident cognitive decline and dementia as outcomes, and other populations from low-resource settings.

## Data Availability

All data used in producing this analysis are available online at https://www.icpsr.umich.edu/web/ICPSR/studies/36633/summary

## Acknowledgements

The Health and Ageing in Africa: Longitudinal Studies in South Africa (HAALSA) study is supported by the U.S. National Institute on Aging of the National Institutes of Health (NIH) (grant number P01 AG041710). The HAALSA-HAALSI study is nested within the SAMRC/Wits University Rural Public Health and Health Transitions Research Unit (Agincourt Health and socio-Demographic Surveillance System), which is supported by the University of the Witwatersrand, Medical Research Council, and Dept of Science and Innovation, South Africa.

## Human Studies statement

The HAALSA protocol was reviewed and approved by the Mpumalanga Provincial Research and Ethics Committee, the University of the Witwatersrand Human Research Ethics Committee (ref. M141159), and the Harvard T.H. Chan School of Public Health Office of Human Research Administration (ref. C13–1608–02). Ethical approval for this secondary data analysis was sought from the Indiana University Institutional Review Board where it was deemed not human subjects research. The study design, execution, and interpretation address heterogeneity in at-risk populations by adding to the limited dementia literature originating from low- and middle-income countries, specifically in populations of sub-Saharan ancestry, who are severely underrepresented in dementia research even as these populations are experiencing rapid growth in dementia burden.

## Consent Statement

Informed consent was obtained from all participants included in the HAALSI study.

## REFERENCES

[1] Nichols E, Steinmetz JD, Vollset SE, Fukutaki K, Chalek J, Abd-Allah F, et al. Estimation of the global prevalence of dementia in 2019 and forecasted prevalence in 2050: An analysis for the global burden of disease study 2019. The Lancet Public Health. 2022;7(2):e105–e25. doi:10.1016/S2468-2667(21)00249-8

[2] Livingston G, Huntley J, Liu KY, Costafreda SG, Selbæk G, Alladi S, et al. Dementia prevention, intervention, and care: 2024 report of the lancet standing commission. Lancet. 2024;404(10452):572-628. doi:10.1016/s0140-6736(24)01296-0

[3] Assuncao N, Sudo FK, Drummond C, de Felice FG, Mattos P. Metabolic syndrome and cognitive decline in the elderly: A systematic review. PLoS One. 2018;13(3):e0194990. doi:10.1371/journal.pone.0194990

[4] Siervo M, Harrison SL, Jagger C, Robinson L, Stephan BC. Metabolic syndrome and longitudinal changes in cognitive function: A systematic review and meta-analysis. J Alzheimers Dis. 2014;41(1):151–61. doi:10.3233/jad-132279

[5] Alberti KG, Eckel RH, Grundy SM, Zimmet PZ, Cleeman JI, Donato KA, et al. Harmonizing the metabolic syndrome: A joint interim statement of the international diabetes federation task force on epidemiology and prevention; national heart, lung, and blood institute; american heart association; world heart federation; international atherosclerosis society; and international association for the study of obesity. Circulation. 2009;120(16):1640–5. doi:10.1161/circulationaha.109.192644

[6] Saklayen M. The global epidemic of the metabolic syndrome. Curr Hypertens Rep. 2018;20(2):12. doi:10.1007/s11906-018-0812-z

[7] Koutsonida M, Markozannes G, Bouras E, Aretouli E, Tsilidis KK. Metabolic syndrome and cognition: A systematic review across cognitive domains and a bibliometric analysis. Front Psychol. 2022;13:981379. doi:10.3389/fpsyg.2022.981379

[8] Rochlani Y, Pothineni NV, Kovelamudi S, Mehta JL. Metabolic syndrome: Pathophysiology, management, and modulation by natural compounds. Therapeutic Advances in Cardiovascular Disease. 2017;11(8):215–25. doi:10.1177/1753944717711379

[9] Kosmas CE, Bousvarou MD, Kostara CE, Papakonstantinou EJ, Salamou E, Guzman E. Insulin resistance and cardiovascular disease. J Int Med Res. 2023;51(3):3000605231164548. doi:10.1177/03000605231164548

[10] Nordestgaard LT, Christoffersen M, Frikke-Schmidt R. Shared risk factors between dementia and atherosclerotic cardiovascular disease. International Journal of Molecular Sciences. 2022;23(17):9777.

[11] Jelinek M, Jurajda M, Duris K. Oxidative stress in the brain: Basic concepts and treatment strategies in stroke. Antioxidants (Basel). 2021;10(12). doi:10.3390/antiox10121886

[12] Kim MY, Kim K, Hong CH, Lee SY, Jung YS. Sex differences in cardiovascular risk factors for dementia. Biomol Ther (Seoul). 2018;26(6):521–32. doi:10.4062/biomolther.2018.159

[13] Tahmi M, Palta P, Luchsinger JA. Metabolic syndrome and cognitive function. Curr Cardiol Rep. 2021;23(12):180. doi:10.1007/s11886-021-01615-y

[14] Alcorn T, Hart E, Smith AE, Feuerriegel D, Stephan BCM, Siervo M, et al. Cross-sectional associations between metabolic syndrome and performance across cognitive domains: A systematic review. Appl Neuropsychol Adult. 2019;26(2):186–99. doi:10.1080/23279095.2017.1363039

[15] Atti AR, Valente S, Iodice A, Caramella I, Ferrari B, Albert U, et al. Metabolic syndrome, mild cognitive impairment, and dementia: A meta-analysis of longitudinal studies. Am J Geriatr Psychiatry. 2019;27(6):625–37. doi:10.1016/j.jagp.2019.01.214

[16] Feng Y, Cheng L, Zhou W, Lu J, Huang H. Metabolic syndrome and the risk of alzheimer’s disease: A meta-analysis. Metab Syndr Relat Disord. 2025;23(1):30–40. doi:10.1089/met.2024.0155

[17] González-Castañeda H, Pineda-García G, Serrano-Medina A, Martínez AL, Bonilla J, Ochoa-Ruíz E. Neuropsychology of metabolic syndrome: A systematic review and meta-analysis. Cogent Psychology. 2021;8(1):1913878. doi:10.1080/23311908.2021.1913878

[18] Pal K, Mukadam N, Petersen I, Cooper C. Mild cognitive impairment and progression to dementia in people with diabetes, prediabetes and metabolic syndrome: A systematic review and meta-analysis. Soc Psychiatry Psychiatr Epidemiol. 2018;53(11):1149–60. doi:10.1007/s00127-018-1581-3

[19] Zuin M, Roncon L, Passaro A, Cervellati C, Zuliani G. Metabolic syndrome and the risk of late onset alzheimer’s disease: An updated review and meta-analysis. Nutr Metab Cardiovasc Dis. 2021;31(8):2244–52. doi:10.1016/j.numecd.2021.03.020

[20] Park Y, Zhu S, Palaniappan L, Heshka S, Carnethon M, Heymsfield S. The metabolic syndrome: Prevalence and associated risk factor findings in the us population from the third national health and nutrition examination survey, 1988-1994. Arch Intern Med. 2003;163(4):427-36. doi:10.1001/archinte.163.4.427

[21] Gyakobo M, Amoah A, Martey-Marbell D, Snow R. Prevalence of the metabolic syndrome in a rural population in ghana. BMC Endocr Disord. 2012;12:25. doi:10.1186/1472-6823-12-25

[22] Motala A, Esterhuizen T, Pirie F, Omar M. The prevalence of metabolic syndrome and determination of the optimal waist circumference cutoff points in a rural south african community. Diabetes Care. 2011;34(4):1032–7. doi:10.2337/dc10-1921

[23] Chen B, Jin X, Guo R, Chen Z, Hou X, Gao F, et al. Metabolic syndrome and cognitive performance among chinese ≥50 years: A cross-sectional study with 3988 participants. Metab Syndr Relat Disord. 2016;14(4):222–7. doi:10.1089/met.2015.0094

[24] Del Brutto OH, Mera RM, Zambrano M. Metabolic syndrome correlates poorly with cognitive performance in stroke-free community-dwelling older adults: A population-based, cross-sectional study in rural ecuador. Aging Clin Exp Res. 2016;28(2):321–5. doi:10.1007/s40520-015-0404-6

[25] Ghosh A, Biswas AK, Banerjee A. A study on cognitive decline with respect to metabolic syndrome and inflammation in elderly indians. Neurol India. 2015;63(4):537–41. doi:10.4103/0028-3886.162037

[26] Madenbay K, Shalkharova Z, Shalkharova Z, Nuskabayeva G, Sadykova K. Association between components of metabolic syndrome and cognitive dysfunction: Cross-sectional study among the population of the turkestan region. Georgian Med News. 2018(278):114–20.

[27] Wang X, Ji L, Tang Z, Ding G, Chen X, Lv J, et al. The association of metabolic syndrome and cognitive impairment in jidong of china: A cross-sectional study. BMC Endocr Disord. 2021;21(1):40. doi:10.1186/s12902-021-00705-w

[28] Wang X, Luan D, Xin S, Liu Y, Gao Q. Association between individual components of metabolic syndrome and cognitive function in northeast rural china. Am J Alzheimers Dis Other Demen. 2019;34(7-8):507–12. doi:10.1177/1533317519865428

[29] Zhan C, Wang Q, Liu J, Wang L, Chen Z, Pang H, et al. Relationship between metabolic syndrome and cognitive function: A population-based study of middle-aged and elderly adults in rural china. Diabetes Metab Syndr Obes. 2021;14:1927–35. doi:10.2147/dmso.S308250

[30] Bahchevanov KM, Dzhambov AM, Chompalov KA, Massaldjieva RI, Atanassova PA, Mitkov MD. Contribution of components of metabolic syndrome to cognitive performance in middle-aged adults. Arch Clin Neuropsychol. 2021;36(4):498–506. doi:10.1093/arclin/acaa081

[31] Foong HF, Hamid TA, Ibrahim R, Haron SA, Shahar S. Chronic condition as a mediator between metabolic syndrome and cognition among community-dwelling older adults: The moderating role of sex. Geriatr Gerontol Int. 2017;17(11):1914–20. doi:10.1111/ggi.12993

[32] García-Lara JM, Aguilar-Navarro S, Gutiérrez-Robledo LM, Avila-Funes JA. The metabolic syndrome, diabetes, and alzheimer’s disease. Rev Invest Clin. 2010;62(4):343–9.

[33] Kopchak OO, Bachyns’ka N, Kholin VO. [cognitive impairment in patients of different age with metabolic syndrome]. Wiad Lek. 2014;67(2 Pt 2):202-6.

[34] Liu M, He Y, Jiang B, Wu L, Wang J, Yang S, et al. Association between metabolic syndrome and mild cognitive impairment and its age difference in a chinese community elderly population. Clin Endocrinol (Oxf). 2015;82(6):844–53. doi:10.1111/cen.12734

[35] Luo L, Yang M, Hao Q, Yue J, Dong B. Cross-sectional study examining the association between metabolic syndrome and cognitive function among the oldest old. J Am Med Dir Assoc. 2013;14(2):105–8. doi:10.1016/j.jamda.2012.10.001

[36] Mehra A, Suri V, Kumari S, Avasthi A, Grover S. Association of mild cognitive impairment and metabolic syndrome in patients with hypertension. Asian J Psychiatr. 2020;53:102185. doi:10.1016/j.ajp.2020.102185

[37] Roriz-Cruz M, Rosset I, Wada T, Sakagami T, Ishine M, De Sá Roriz-Filho J, et al. Cognitive impairment and frontal-subcortical geriatric syndrome are associated with metabolic syndrome in a stroke-free population. Neurobiol Aging. 2007;28(11):1723–36. doi:10.1016/j.neurobiolaging.2006.07.013

[38] Zheng W, Zhou X, Yin J, Liu H, Yin W, Zhang W, et al. Metabolic syndrome-related cognitive impairment with white matter hyperintensities and functional network analysis. Obesity (Silver Spring). 2023;31(10):2557–67. doi:10.1002/oby.23873

[39] Liang H, Fang Y. Association of metabolic syndrome and its components with motoric cognitive risk syndrome among older adults: A population-based prospective longitudinal cohort study. J Psychosom Res. 2023;173:111456. doi:10.1016/j.jpsychores.2023.111456

[40] Liu Y, Zang B, Shao J, Ning N, He L, Ma Y. Predictor of cognitive impairment: Metabolic syndrome or circadian syndrome. BMC Geriatr. 2023;23(1):408. doi:10.1186/s12877-023-03996-x

[41] Shigaeff N, Amaro E, Franco FGM, Jacinto AF, Chiochetta G, Cendoroglo MS, et al. Functional magnetic resonance imaging response as an early biomarker of cognitive decline in elderly patients with metabolic syndrome. Arch Gerontol Geriatr. 2017;73:1–7. doi:10.1016/j.archger.2017.07.002

[42] Tian YM, Zhang WS, Jiang CQ, Zhu F, Jin YL, Yeung SLA, et al. Longitudinal association of changes in metabolic syndrome with cognitive function: 12-year follow-up of the guangzhou biobank cohort study. Diabetes Metab J. 2025;49(1):60–79. doi:10.4093/dmj.2024.0117

[43] Bowo-Ngandji A, Kenmoe S, Ebogo-Belobo J, Kenfack-Momo R, Takuissu G, Kengne-Ndé C, et al. Prevalence of the metabolic syndrome in african populations: A systematic review and meta-analysis. PLoS One. 2023;18(7):e0289155. doi:10.1371/journal.pone.0289155

[44] Charles-Davies M, Ajayi O. Redefining the metabolic syndrome in africa: A systematic review between 2005 and 2022. Dubai Diabetes and Endocrinology Journal. 2023;29(2):89–98. doi:10.1159/000531552

[45] Crowther N, Norris S. The current waist circumference cut point used for the diagnosis of metabolic syndrome in sub-saharan african women is not appropriate. PLOS ONE. 2012;7(11):e48883. doi:10.1371/journal.pone.0048883

[46] Ekoru K, Murphy GAV, Young EH, Delisle H, Jerome CS, Assah F, et al. Deriving an optimal threshold of waist circumference for detecting cardiometabolic risk in sub-saharan africa. Int J Obes (Lond). 2017;42(3):487–94. doi:10.1038/ijo.2017.240

[47] Gradidge PJ, Norris SA, Crowther NJ. The effect of obesity on the waist circumference cut-point used for the diagnosis of the metabolic syndrome in african women: Results from the sweet study. Int J Environ Res Public Health. 2022;19(16). doi:10.3390/ijerph191610250

[48] Hoebel S, Malan L, De Ridder JH. Determining ethnic-, gender-, and age-specific waist circumference cut-off points to predict metabolic syndrome: The sympathetic activity and ambulatory blood pressure in africans (sabpa) study. Journal of Endocrinology, Metabolism and Diabetes of South Africa. 2013;18(2):88–96. doi:10.1080/22201009.2013.10872311

[49] Owolabi EO, Ter Goon D, Adeniyi OV, Ajayi AI. Optimal waist circumference cut-off points for predicting metabolic syndrome among low-income black south african adults. BMC Research Notes. 2018;11(1):22. doi:10.1186/s13104-018-3136-9

[50] Prinsloo J, Malan L, de Ridder JH, Potgieter JC, Steyn HS. Determining the waist circumference cut off which best predicts the metabolic syndrome components in urban africans: The sabpa study. Exp Clin Endocrinol Diabetes. 2011;119(10):599–603. doi:10.1055/s-0031-1280801

[51] Berkman LF. Health and aging in africa: A longitudinal study of an indepth community in south africa [haalsi]: Agincourt, south africa, 2015-2022. Inter-university Consortium for Political and Social Research [distributor]; 2023.

[52] Gómez-Olivé F, Montana L, Wagner R, Kabudula C, Rohr J, Kahn K, et al. Cohort profile: Health and ageing in africa: A longitudinal study of an indepth community in south africa (haalsi). Int J Epidemiol. 2018;47(3):689–90j. doi:10.1093/ije/dyx247

[53] Kahn K, Collinson MA, Gómez-Olivé FX, Mokoena O, Twine R, Mee P, et al. Profile: Agincourt health and socio-demographic surveillance system. Int J Epidemiol. 2012;41(4):988–1001. doi:10.1093/ije/dys115

[54] Kallaway P. Apartheid and education: The education of black south africans: Ravan Press; 1984.

[55] United nations. Report of the special committee on the policies of apartheid of the government of the republic of south africa. 1963.

[56] Tabák AG, Herder C, Rathmann W, Brunner EJ, Kivimäki M. Prediabetes: A high-risk state for diabetes development. Lancet. 2012;379(9833):2279-90. doi:10.1016/s0140-6736(12)60283-9

[57] Erasmus RT, Soita DJ, Hassan MS, Blanco-Blanco E, Vergotine Z, Kegne AP, et al. High prevalence of diabetes mellitus and metabolic syndrome in a south african coloured population: Baseline data of a study in bellville, cape town. S Afr Med J. 2012;102(11 Pt 1):841-4. doi:10.7196/samj.5670

[58] Gross AL, Power MC, Albert MS, Deal JA, Gottesman RF, Griswold M, et al. Application of latent variable methods to the study of cognitive decline when tests change over time. Epidemiology. 2015;26(6):878–87. doi:10.1097/ede.0000000000000379

[59] Chakraborty R, Kobayashi LC, Jock J, Wing C, Chen X, Phillips M, et al. Child support grant expansion and cognitive function among women in rural south africa: Findings from a natural experiment in the haalsi cohort. PLoS One. 2024;19(3):e0297673. doi:10.1371/journal.pone.0297673

[60] Jock J, Kobayashi L, Chakraborty R, Chen X, Wing C, Berkman L, et al. Effects of pension eligibility expansion on men’s cognitive function: Findings from rural south africa. Journal of Aging & Social Policy.1–20. doi:10.1080/08959420.2023.2195785

[61] Weuve J, Proust-Lima C, Power MC, Gross AL, Hofer SM, Thiébaut R, et al. Guidelines for reporting methodological challenges and evaluating potential bias in dementia research. Alzheimers Dement. 2015;11(9):1098–109. doi:10.1016/j.jalz.2015.06.1885

[62] R Core Team. R: A language and environment for statistical computing. R Foundation for Statistical Computing. Vienna, Austria.: R Core Team; 2024.

[63] Bates DM, Martin; Bolker, Ben; Walker, Steve. Fitting linear mixed-effects models using lme4. Journal of Statistical Software. 2015;67(1):1—48. doi:10.18637/jss.v067.i01

[64] Pant V, Gautam K, Pradhan S. Postprandial blood glucose can be less than fasting blood glucose and this is not a laboratory error. JNMA J Nepal Med Assoc. 2019;57(215):67–8. doi:10.31729/jnma.3985

[65] Gallegos D. Effects of food and nutrition insecurity on global health. N Engl J Med. 2025;392(7):686–97. doi:10.1056/NEJMra2406458

[66] Park S, Strauss S. Food insecurity as a predictor of metabolic syndrome in u.S. Female adults. Public Health Nursing. 2020;37(5):663–70. doi:10.1111/phn.12781

[67] Peltzer K, Phaswana-Mafuya N. Fruit and vegetable intake and associated factors in older adults in south africa. Glob Health Action. 2012;5:1–8. doi:10.3402/gha.v5i0.18668

[68] Gradidge P, Crowther N. Review: Metabolic syndrome in black south african women. Ethn Dis. 2017;27(2):189–200. doi:10.18865/ed.27.2.189

[69] Marden JR, Tchetgen Tchetgen EJ, Kawachi I, Glymour MM. Contribution of socioeconomic status at 3 life-course periods to late-life memory function and decline: Early and late predictors of dementia risk. Am J Epidemiol. 2017;186(7):805–14. doi:10.1093/aje/kwx155

[70] Harvey PD. Domains of cognition and their assessment. Dialogues Clin Neurosci. 2019;21(3):227–37. doi:10.31887/DCNS.2019.21.3/pharvey

[71] Booncharoen K, Hiransuthikul A, Thanapornsangsuth P, Thanprasertsuk S, Sarutikriangkri Y, Tangnimitchoke S, et al. Episodic memory decline symptoms are strong predictors of alzheimer’s disease defined by positron emission tomography in participants with amnestic mild cognitive impairment and mild dementia. Alzheimer’s & Dementia. 2023;19(S18):e073005. doi:10.1002/alz.073005

[72] Tromp D, Dufour A, Lithfous S, Pebayle T, Després O. Episodic memory in normal aging and alzheimer disease: Insights from imaging and behavioral studies. Ageing Research Reviews. 2015;24:232–62. doi:10.1016/j.arr.2015.08.006

