## Supplementary Table 1 for "Metabolic syndrome and memory decline: evidence from a longitudinal aging cohort in rural South Africa"

Supplementary Table 1. Adjusted* effect of MetS component conditions on memory scores: HAALSA Indepth participants, 2014/15-2021/22

### Difference in baseline memory score

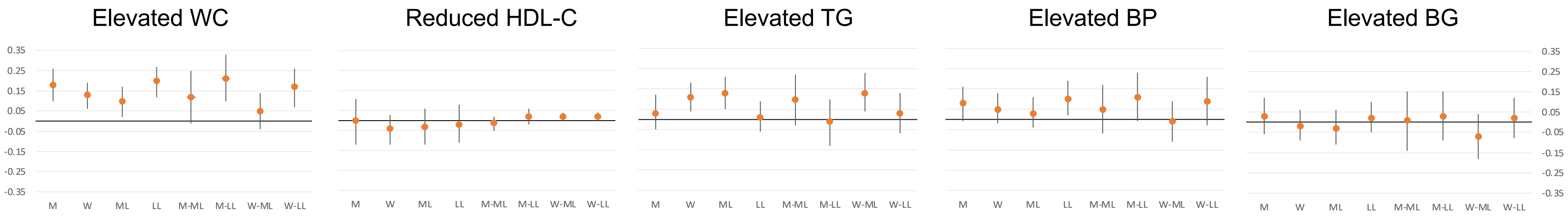


### Difference in rate of memory decline

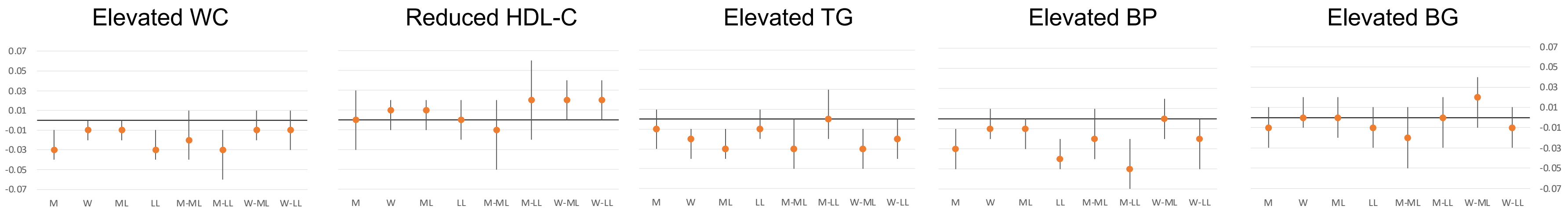


*Adjusted for sex, continuous age, having children, literacy, education level, consumption quintile, marital status, self-reported HIV status, and country of birth.
WC = waist circumference; HDL-C = high density lipoprotein cholesterol; TG = triglyceride; BP = blood pressure; BG = blood glucose
M = men; W = women; ML = mid-life; LL = later-life
Note: Effect estimate in standard deviation units.
