## Supplementary Table 2 for "Metabolic syndrome and memory decline: evidence from a longitudinal aging cohort in rural South Africa"

Supplementary Table 2. Sensitivity analysis using Europid waist circumference thresholds

Association between MetS and memory scores

| | Baseline memory score | | | Memory score decline | | |
|---|---|---|---|---|---|---|
| | β | 95% CI | *P* value | β | 95% CI | *P* value |
| Full population | | | | | | |
| Unadjusted | **0.07** | **(0.01, 0.13)** | **0.015** | **-0.02** | **(-0.03, -0.01)** | **0.003** |
| Adjusted* | **0.08** | **(0.03, 0.14)** | **0.003** | **-0.02** | **(-0.03, 0.00)** | **0.005** |
| By Sex† | | | | | | |
| Men | 0.09 | (0.00, 0.18) | 0.056 | -0.01 | (-0.03, 0.01) | 0.145 |
| Women | 0.06 | (-0.01, 0.13) | 0.071 | -0.01 | (-0.03, 0.00) | 0.056 |
| By Age* | | | | | | |
| Mid-life | **0.09** | **(0.01, 0.17)** | **0.021** | **-0.02** | **(-0.03, 0.00)** | **0.035** |
| Later-life | 0.06 | (-0.02, 0.13) | 0.131 | -0.02 | (-0.03, 0.00) | 0.061 |
| By Sex and Age† | | | | | | |
| Mid-life men | 0.08 | (-0.06, 0.22) | 0.247 | -0.02 | (-0.05, 0.01) | 0.161 |
| Later-life men | 0.09 | (-0.03, 0.21) | 0.129 | -0.01 | (-0.04, 0.02) | 0.463 |
| Mid-life women | 0.08 | (-0.02, 0.17) | 0.110 | -0.01 | (-0.03, 0.00) | 0.148 |
| Later-life women | 0.01 | (-0.09, 0.11) | 0.827 | -0.01 | (-0.04, 0.01) | 0.186 |

Association between elevated waist circumference and memory scores

| | Baseline memory score | | | Memory score decline | | |
|---|---|---|---|---|---|---|
| | β | 95% CI | *P* value | β | 95% CI | *P* value |
| Full population | | | | | | |
| Unadjusted | **0.17** | **(0.11, 0.23)** | **<0.001** | **-0.02** | **(-0.03, -0.01)** | **<0.001** |
| Adjusted* | **0.17** | **(0.11, 0.22)** | **<0.001** | **-0.02** | **(-0.03, -0.01)** | **<0.001** |
| By Sex† | | | | | | |
| Men | **0.20** | **(0.12, 0.28)** | **<0.001** | **-0.03** | **(-0.05, -0.01)** | **0.002** |
| Women | **0.12** | **(0.03, 0.21)** | **0.009** | -0.01 | (-0.03, 0.01) | 0.303 |
| By Age* | | | | | | |
| Mid-life | **0.15** | **(0.07, 0.23)** | **<0.001** | -0.01 | (-0.03, 0.00) | 0.061 |
| Later-life | **0.17** | **(0.09, 0.25)** | **<0.001** | **-0.03** | **(-0.04, -0.010** | **0.002** |
| By Sex and Age† | | | | | | |
| Mid-life men | **0.15** | **(0.03, 0.28)** | **0.018** | -0.02 | (-0.05, 0.01) | 0.123 |
| Later-life men | **0.23** | **(0.12, 0.34)** | **<0.001** | **-0.04** | **(-0.06, -0.01)** | **0.004** |
| Mid-life women | **0.13** | **(0.00, 0.25)** | **0.043** | -0.01 | (-0.04, 0.01) | 0.358 |
| Later-life women | 0.07 | (-0.06, 0.20) | 0.309 | -0.01 | (-0.03, 0.02) | 0.691 |

* Adjusted for sex, continuous age, having children, literacy, education level, consumption quintile, marital status, self-reported HIV status, and country of birth.

†Adjusted for continuous age, having children, literacy, education level, consumption quintile, marital status, self-reported HIV status, and country of birth.

Note: Effect estimate in standard deviation units. Elevated waist circumference using Europid thresholds is >= 94.0 cm for men and >= 80.0 cm for women
