## Supplementary Table 3 for "Metabolic syndrome and memory decline: evidence from a longitudinal aging cohort in rural South Africa"

Supplementary Table 3. Sensitivity analysis excluding those with missing fasting data from determination of elevated blood glucose

### Association between MetS and memory scores

| | Baseline memory score | | | Memory score decline | | |
|---|---|---|---|---|---|---|
| | β | 95% CI | *P* value | β | 95% CI | *P* value |
| Full population | | | | | | |
| Unadjusted | **0.08** | **(0.02, 0.14)** | **0.01** | **-0.01** | **(-0.02, 0.00)** | **0.021** |
| Adjusted* | **0.07** | **(0.01, 0.13)** | **0.013** | **-0.01** | **(-0.02, 0.00)** | **0.039** |
| By Sex† | | | | | | |
| Men | 0.07 | (-0.02, 0.16) | 0.150 | -0.01 | (-0.03, 0.01) | 0.376 |
| Women | 0.06 | (-0.01, 0.13) | 0.083 | -0.01 | (-0.03, 0.00) | 0.109 |
| By Age* | | | | | | |
| Mid-life | 0.07 | (-0.01, 0.15) | 0.101 | -0.01 | (-0.03, 0.00) | 0.116 |
| Later-life | 0.06 | (-0.01, 0.03) | 0.454 | -0.01 | (-0.03, 0.01) | 0.183 |
| By Sex and Age† | | | | | | |
| Mid-life men | 0.08 | (-0.07, 0.22) | 0.291 | -0.02 | (-0.05, 0.01) | 0.235 |
| Later-life men | 0.06 | (-0.06, 0.18) | 0.322 | 0.00 | (-0.03, 0.03) | 0.864 |
| Mid-life women | 0.04 | (-0.06, 0.14) | 0.424 | -0.01 | (-0.03, 0.01) | 0.364 |
| Later-life women | 0.04 | (-0.06, 0.14) | 0.403 | -0.01 | (-0.04, 0.01) | 0.177 |

### Association between elevated blood glucose and memory scores

| | Baseline memory score | | | Memory score decline | | |
|---|---|---|---|---|---|---|
| | β | 95% CI | *P* value | β | 95% CI | *P* value |
| Full population | | | | | | |
| Unadjusted | **-0.08** | **(-0.14, -0.01)** | **0.021** | 0.00 | (-0.01, 0.01) | 0.923 |
| Adjusted[1] | -0.01 | (-0.06, 0.05) | 0.807 | 0.00 | (-0.01, 0.01) | 0.774 |
| By Sex[2] | | | | | | |
| Men | 0.01 | (-0.08, 0.10) | 0.822 | -0.01 | (-0.03, 0.01) | 0.379 |
| Women | -0.02 | (-0.10, 0.05) | 0.574 | 0.01 | (-0.01, 0.02) | 0.498 |
| By Age[1] | | | | | | |
| Mid-life | -0.04 | (-0.13, 0.05) | 0.357 | 0.00 | (-0.01, 0.02) | 0.673 |
| Later-life | 0.01 | (-0.01, 0.03) | 0.419 | -0.01 | (-0.02, 0.01) | 0.410 |
| By Sex and Age[2] | | | | | | |
| Mid-life men | -0.02 | (-0.17, 0.13) | 0.782 | -0.02 | (-0.05, 0.01) | 0.260 |
| Later-life men | 0.02 | (-0.10, 0.14) | 0.752 | 0.00 | (-0.03, 0.02) | 0.829 |
| Mid-life women | -0.08 | (-0.18, 0.03) | 0.168 | 0.02 | (0.00, 0.04) | 0.107 |
| Later-life women | 0.01 | (-0.09, 0.11) | 0.822 | -0.01 | (-0.03, 0.01) | 0.502 |

*Adjusted for sex, continuous age, having children, literacy, education level, consumption quintile, marital status, self-reported HIV status, and country of birth.

[†]Adjusted for continuous age, having children, literacy, education level, consumption quintile, marital status, self-reported HIV status, and country of birth.

Note: Effect estimate in standard deviation units. 52 participants had missing fasting data, so an additional 52 participants had elevated blood glucose assigned as NA in MetS determination.
